# Identifying DNA Methylation Patterns in Post COVID-19 Condition: Insights from a One-Year Prospective Cohort Study

**DOI:** 10.1101/2025.02.28.25323075

**Authors:** Christoffer Granvik, Jelena Smiljanić, Alicia Lind, David Martínez-Enguita, Shumaila Sayyab, Clas Ahlm, Mattias N. E. Forsell, Sara Cajander, Mika Gustafsson, Martin Rosvall, Johan Normark, Maria Lerm

**Author notes:** shared first authorship. shared last authorship (Martin Rosvall, Johan Normark, Maria Lerm).

## Abstract

The mechanisms underlying Post COVID-19 condition (PCC), with its range of long-lasting symptoms, remain unclear. This study investigates DNA methylation patterns over one year in a subset of non-hospitalized COVID-19 patients with persistent symptoms and reduced quality of life, termed PCC+ (Post COVID-19 condition plus). In a cohort of 22 PCC+ individuals and 22 matched COVID-19 convalescents (PCC-), we identified distinct DNA methylation differences between the groups that diminish over time. Methylation changes in the TXNRD1 gene were significantly associated with cognitive symptoms and fatigue, implicating redox imbalance in PCC pathology. Pathway analysis revealed enrichment in PI3K-Akt and AMPK signaling pathways, potentially underlying the observed efficacy of metformin in reducing PCC incidence. While we found no differences in epigenetic age acceleration between the groups, we observed longitudinal changes in the methylation of the RAS and RAP1 signaling pathways. These findings provide crucial insights into PCC+ mechanisms and suggest oxidative stress pathways as promising targets for therapeutic interventions.

## 1 Introduction

A growing body of research has documented the long-lasting clinical effects following COVID-19 infection, observed in both hospitalized and non-hospitalized patients [1]. These sequelae following COVID-19 infection are referred to by various terms, including long COVID, Post-Acute COVID-19 Syndrome (PACS), and chronic COVID. The persistence of symptoms beyond three months, defined by the WHO as Post COVID-19 Condition (PCC) [2], varies significantly in presentation and severity among affected individuals [3]. This symptom-based definition poses challenges for research, as many COVID-19 patients experience some degree of persisting symptoms. No established outcome criteria or measures exist to predict PCC prognosis, track disease progression, assess long-term outcomes, or address its lasting impacts.

Among individuals with persistent symptoms, a distinct subset also suffers from reduced health-related quality of life after infection [4]. Referred to as Post COVID-19 condition plus (PCC+), this subset highlights the complex spectrum of outcomes following COVID-19. The refined definition addresses limitations in the broader WHO definition of PCC, which is often too nonspecific to effectively identify patients requiring assistance. However, the biological mechanisms that distinguish PCC+ from other post-viral conditions and COVID-19 convalescents (PCC-) remain unclear.

To better understand these mechanisms, efforts have focused on identifying biological signatures that differentiate individuals with various persistent sequelae from those who fully recover [5, 6]. Epigenetic modifications through DNA methylation (DNAm) offer a promising avenue for understanding these long-term effects, as they reflect both genetic predispositions and environmental influences, including viral infections [7, 8]. Analysis of epigenetic biosignatures in peripheral blood mononuclear cells suggested a unique DNAm signature in patients with PCC, differentiating them from both healthy COVID-19 convalescents and controls [9]. Other studies have identified changes in the innate immune response to viral infections [10], disruptions in circadian rhythm-regulating pathways [11], and evidence of accelerated biological aging in individuals with PCC [12]. However, the focus on single time points and lack of longitudinal design in these studies limits their ability to capture the progression and dynamics of these changes over time.

We analyzed longitudinal DNAm changes in blood immune cells to investigate potential epigenetic contributions to post-viral sequelae in non-hospitalized individuals following COVID-19 infection. We found specific DNAm patterns in peripheral blood mononuclear cells (PBMCs) that distinguish PCC+ individuals from COVID-19 convalescents. These methylation differences, associated with redox imbalance, diminished over time, reflecting gradual symptom improvement in many PCC+ patients. Enriched pathways suggest potential therapeutic targets and provide insights into mechanisms that may help reduce PCC incidence.

## 2 Results

### 2.1 Characteristics of the study population and symptoms at follow-up

Individuals with initial mild COVID-19 infection and PCC+ at follow-up were selected and COVID-19 convalescents (PCC-) as matched controls based on initial infection severity, age, sex, and, BMI. PCC- was defined as individuals who did not fulfil the criteria for PCC+ with persistent symptoms and reduced health-related quality of life. We included 22 individuals with PCC+ and 22 PCC- (Fig. 1). There were no significant differences in comorbidities, education, or smoking status between the PCC+ and PCC- (Table 1). At follow-up, there were five missing samples at 3 months, one at 6 months and no missing samples at 12 months. The missing samples introduced no significant differences in baseline characteristics between the groups.

**Figure 1:**
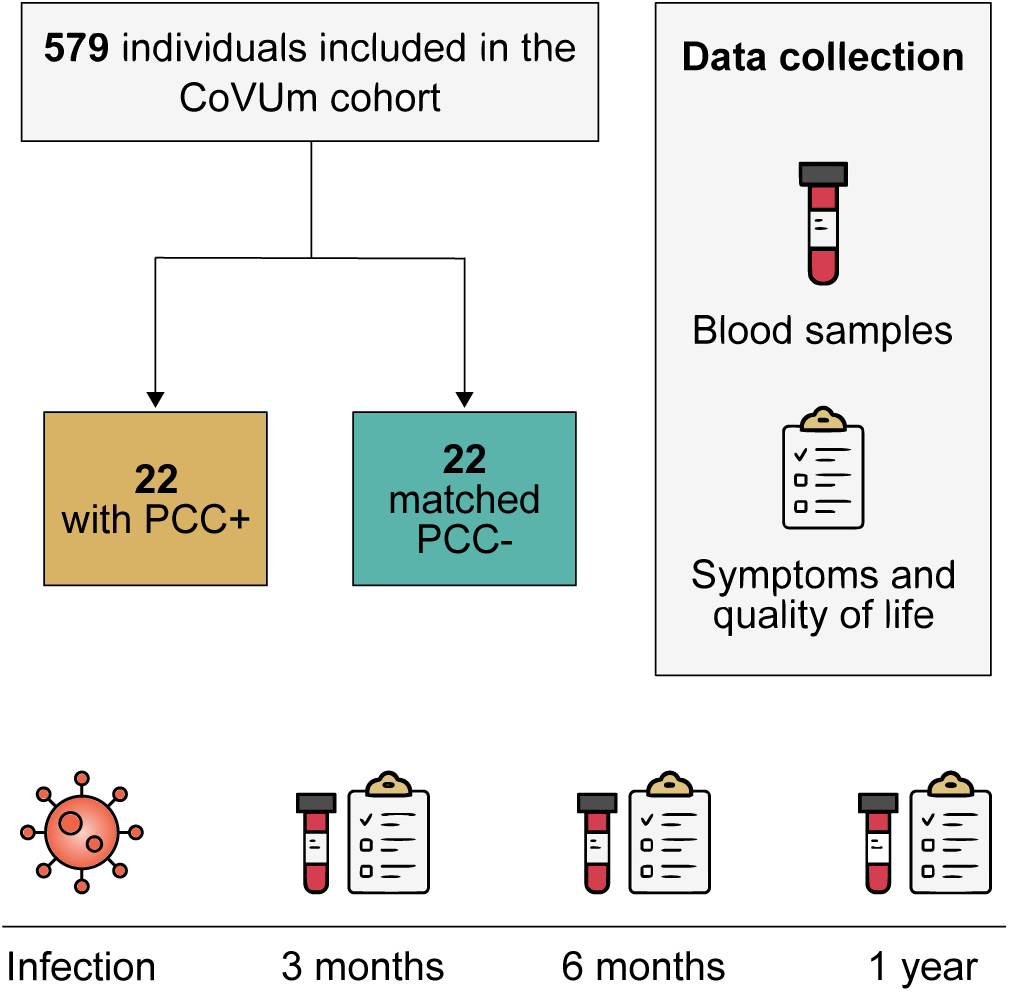
Study design. Individuals with initial mild COVID-19 infection and post COVID-19 condi- tion plus (PCC+, *N* = 22) were selected for methylation analysis. COVID-19 convalescents (PCC-) were matched by age, sex, and BMI (*N* = 22).

**Table 1:**
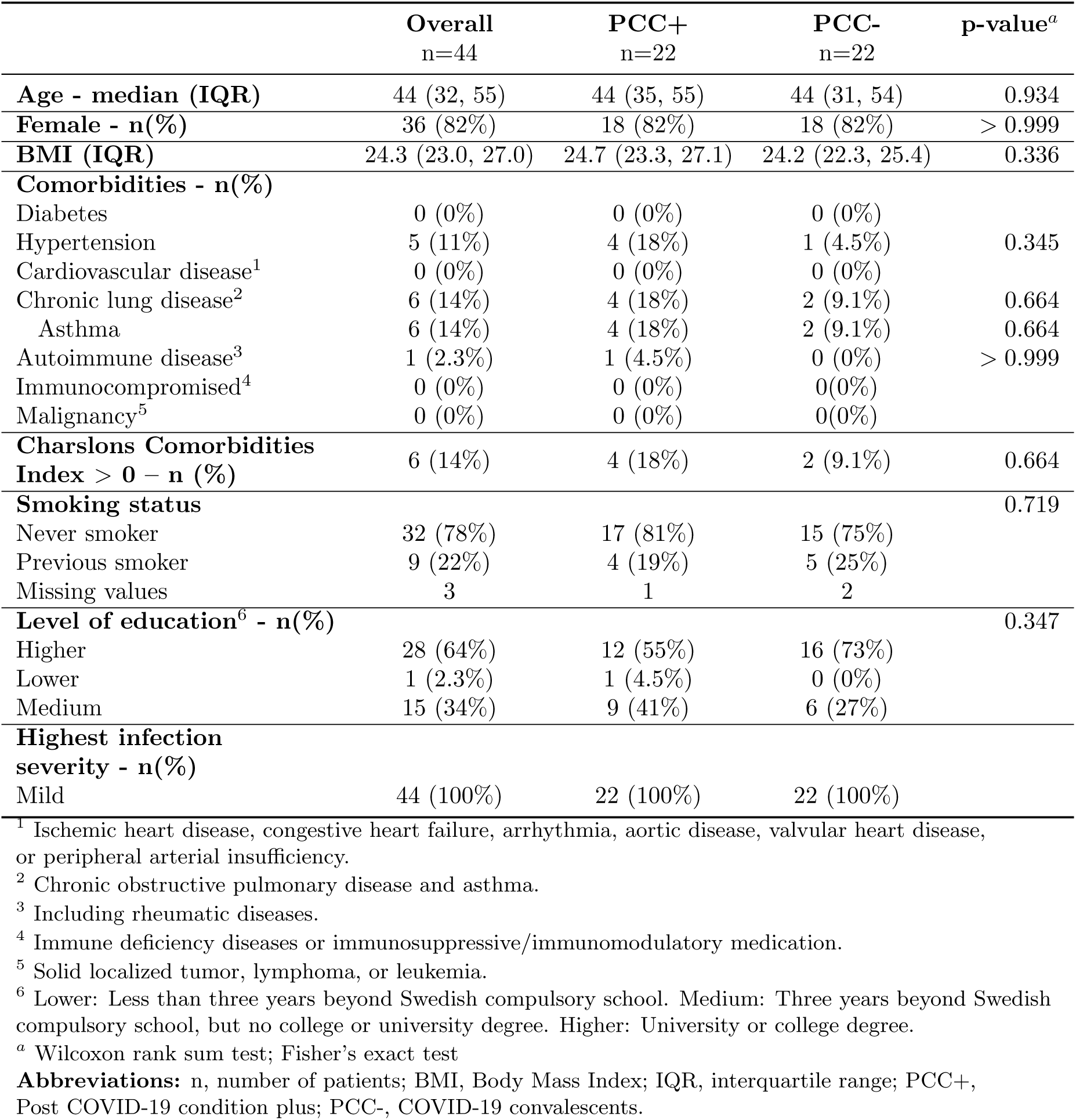
Demographics and baseline characteristics of the study cohort, divided into PCC+ and PCC-.

To account for potential variations in infection severity among non-hospitalized individuals, we analyzed clinical chemistry data from the first 30 days after infection onset using principal component analysis. No significant differences were observed between individuals with PCC+ and PCC- (Supplementary Fig. A1).

The PCC+ group showed a high prevalence of persisting symptoms at follow-ups. The most common were neurological and psychiatric symptoms such as difficulties finding words, mental fatigue, concentration difficulties, and memory difficulties. The number of symptoms decreased over time as PCC+ individuals gradually recovered. An additional follow-up at 24 months showed even further improvement. The prevalence of symptoms in each group is presented in Table 2.

**Table 2:**
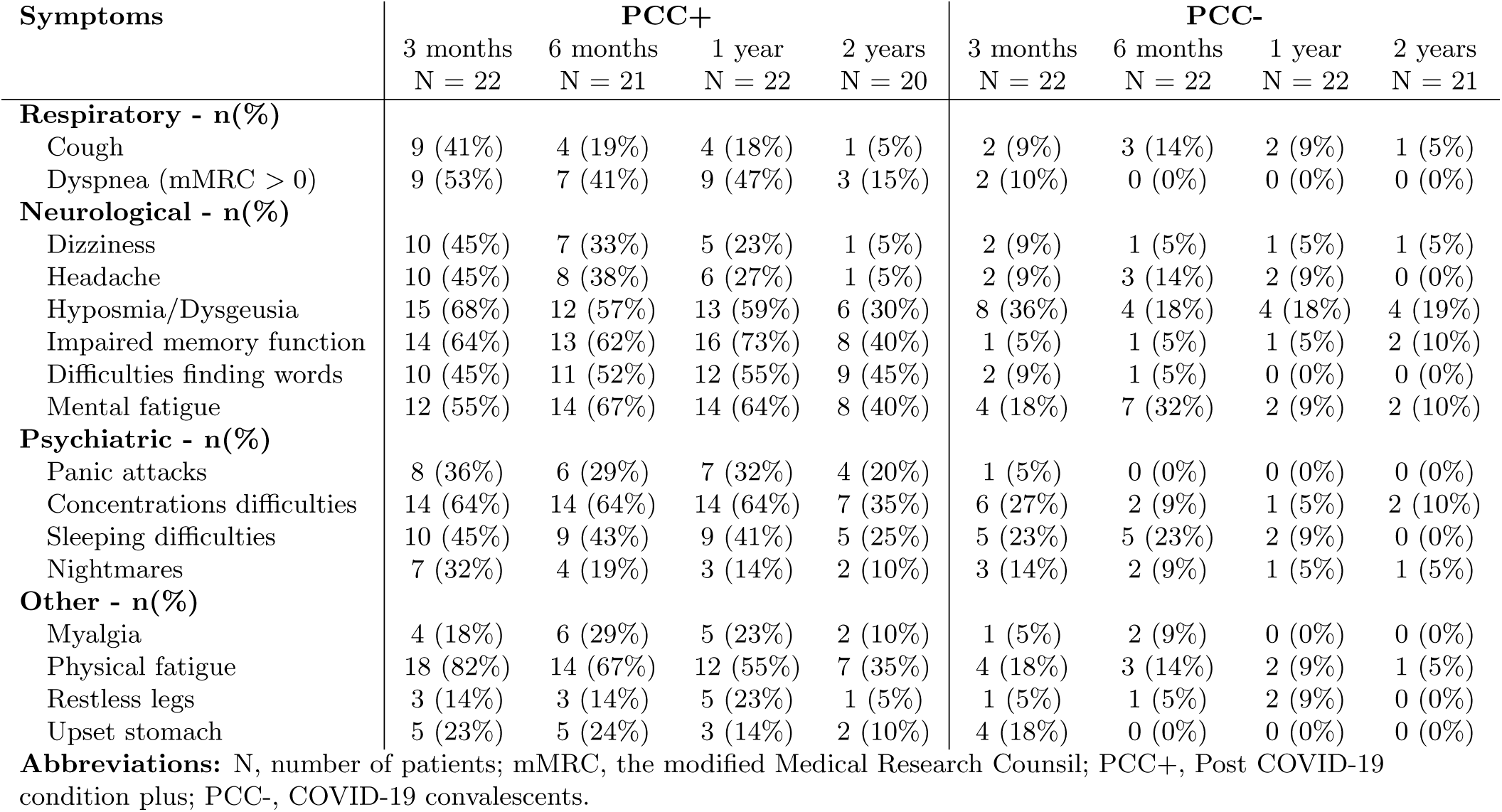
Reported symptoms at follow-up, divided in PCC+ and PCC-.

| Symptoms | PCC+ |  |  |  | PCC- |  |  |  |
| --- | --- | --- | --- | --- | --- | --- | --- | --- |
|  | 3 months<br>N = 22 | 6 months<br>N = 21 | 1 year<br>N = 22 | 2 years<br>N = 20 | 3 months<br>N = 22 | 6 months<br>N = 22 | 1 year<br>N = 22 | 2 years<br>N = 21 |
| **Respiratory - n(%)** |  |  |  |  |  |  |  |  |
| Cough | 9 (41%) | 4 (19%) | 4 (18%) | 1 (5%) | 2 (9%) | 3 (14%) | 2 (9%) | 1 (5%) |
| Dyspnea (mMRC > 0) | 9 (53%) | 7 (41%) | 9 (47%) | 3 (15%) | 2 (10%) | 0 (0%) | 0 (0%) | 0 (0%) |
| **Neurological - n(%)** |  |  |  |  |  |  |  |  |
| Dizziness | 10 (45%) | 7 (33%) | 5 (23%) | 1 (5%) | 2 (9%) | 1 (5%) | 1 (5%) | 1 (5%) |
| Headache | 10 (45%) | 8 (38%) | 6 (27%) | 1 (5%) | 2 (9%) | 3 (14%) | 2 (9%) | 0 (0%) |
| Hyposmia/Dysgeusia | 15 (68%) | 12 (57%) | 13 (59%) | 6 (30%) | 8 (36%) | 4 (18%) | 4 (18%) | 4 (19%) |
| Impaired memory function | 14 (64%) | 13 (62%) | 16 (73%) | 8 (40%) | 1 (5%) | 1 (5%) | 1 (5%) | 2 (10%) |
| Difficulties finding words | 10 (45%) | 11 (52%) | 12 (55%) | 9 (45%) | 2 (9%) | 1 (5%) | 0 (0%) | 0 (0%) |
| Mental fatigue | 12 (55%) | 14 (67%) | 14 (64%) | 8 (40%) | 4 (18%) | 7 (32%) | 2 (9%) | 2 (10%) |
| **Psychiatric - n(%)** |  |  |  |  |  |  |  |  |
| Panic attacks | 8 (36%) | 6 (29%) | 7 (32%) | 4 (20%) | 1 (5%) | 0 (0%) | 0 (0%) | 0 (0%) |
| Concentrations difficulties | 14 (64%) | 14 (64%) | 14 (64%) | 7 (35%) | 6 (27%) | 2 (9%) | 1 (5%) | 2 (10%) |
| Sleeping difficulties | 10 (45%) | 9 (43%) | 9 (41%) | 5 (25%) | 5 (23%) | 5 (23%) | 2 (9%) | 0 (0%) |
| Nightmares | 7 (32%) | 4 (19%) | 3 (14%) | 2 (10%) | 3 (14%) | 2 (9%) | 1 (5%) | 1 (5%) |
| **Other - n(%)** |  |  |  |  |  |  |  |  |
| Myalgia | 4 (18%) | 6 (29%) | 5 (23%) | 2 (10%) | 1 (5%) | 2 (9%) | 0 (0%) | 0 (0%) |
| Physical fatigue | 18 (82%) | 14 (67%) | 12 (55%) | 7 (35%) | 4 (18%) | 3 (14%) | 2 (9%) | 1 (5%) |
| Restless legs | 3 (14%) | 3 (14%) | 5 (23%) | 1 (5%) | 1 (5%) | 1 (5%) | 2 (9%) | 0 (0%) |
| Upset stomach | 5 (23%) | 5 (24%) | 3 (14%) | 2 (10%) | 4 (18%) | 0 (0%) | 0 (0%) | 0 (0%) |

**Abbreviations:** N, number of patients; mMRC, the modified Medical Research Council; PCC+, Post COVID-19 condition plus; PCC-, COVID-19 convalescents.

### 2.2 Thioredoxin reductase 1 (TXNRD1) is differentially methylated in individuals with PCC+ and is associated with cognitive symptoms and fatigue

We sought to identify relevant differentially methylated CpG sites (DMCs), comparing PCC+ with PCC- at each time point to better understand the pathology behind PCC+. At 3 months, we observed 16521 DMCs with a nominal p-value *<* 0.01. The number of DMCs decreased over time, with 6736 identified at 6 months and 4062 at 12 months (Fig. 2a). After annotating DMCs to genes (DMGs), we identified 4017 DMGs separating PCC+ from PCC- at 3 months, 2019 at 6 months, and 928 at 1 year. KEGG pathway analysis of DMGs at each time point showed enrichment of several signaling pathways (Fig. 2c). The most enriched pathway was the PI3K-Akt signaling pathway at 3 months with 167 enriched genes. Gene Ontology enrichment analysis revealed that multiple genes were linked to the regulation of trans-synaptic signaling and modulation of chemical synaptic transmission at 3 months after infection (Fig. A3).

**Figure 2:**
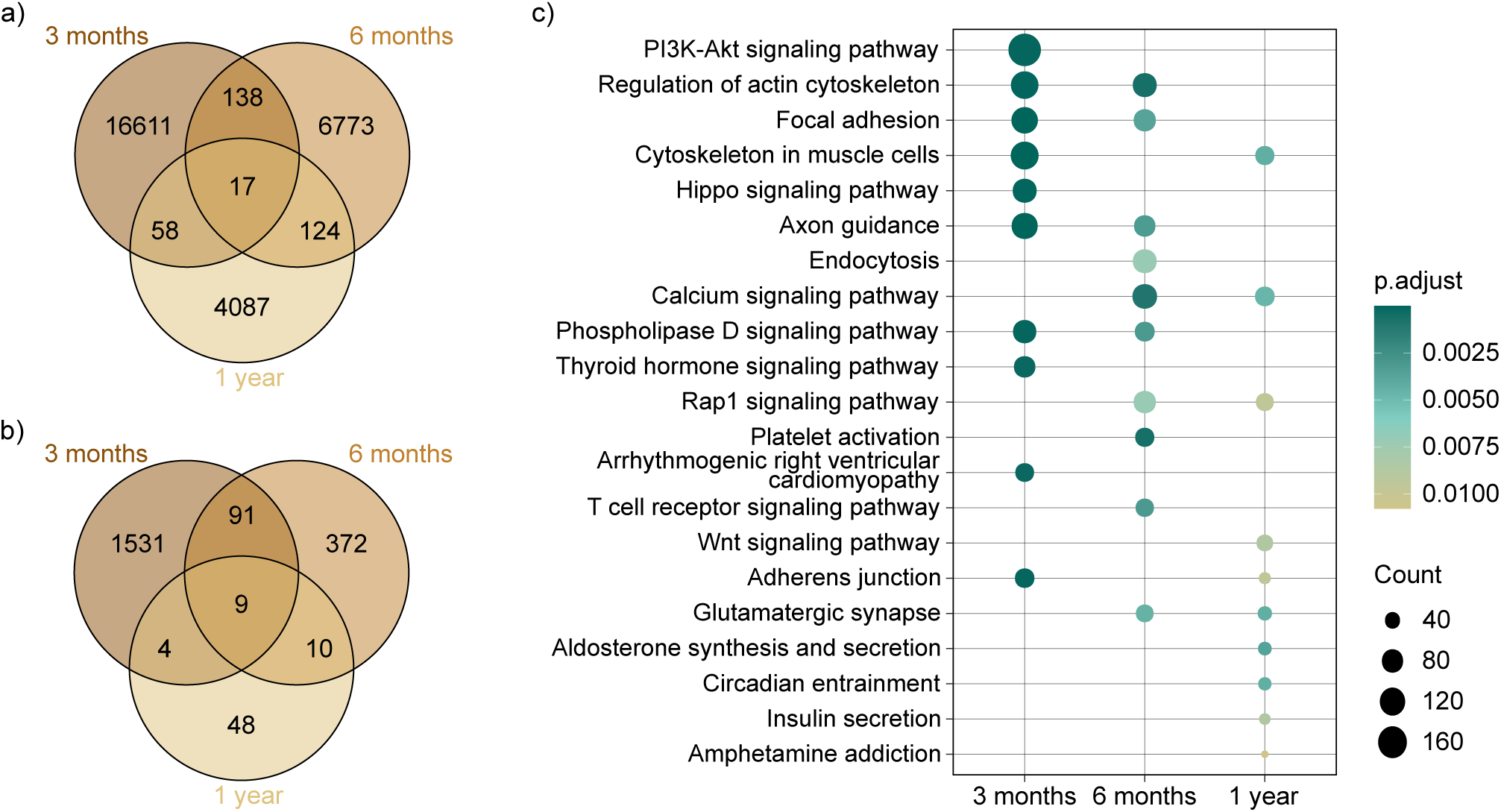
Differential methylation analysis. Venn diagrams showing the overlap of a) differentially methylated CpGs (DMCs) and b) genes in differentially methylated regions (DMRs) between PCC+ and PCC- groups at 3 months, 6 months, and 12 months. The findings suggest that most DNA methylation changes are short-term. However, CpGs and regions displaying consistent methylation changes across all three time points are associated with the genes CPLX1, EID3, and TXNRD1. c) KEGG pathway enrichment analysis of differentially methylated genes (DMGs) between PCC+ and PCC- samples. The horizontal axis represents different time points, while the vertical axis lists the top 10 enriched KEGG pathways. Node size indicates the number of DMGs associated with each pathway, and node color intensity corresponds to the adjusted p-value, with dark green colors reflecting more significant enrichment.

**Figure 3:**
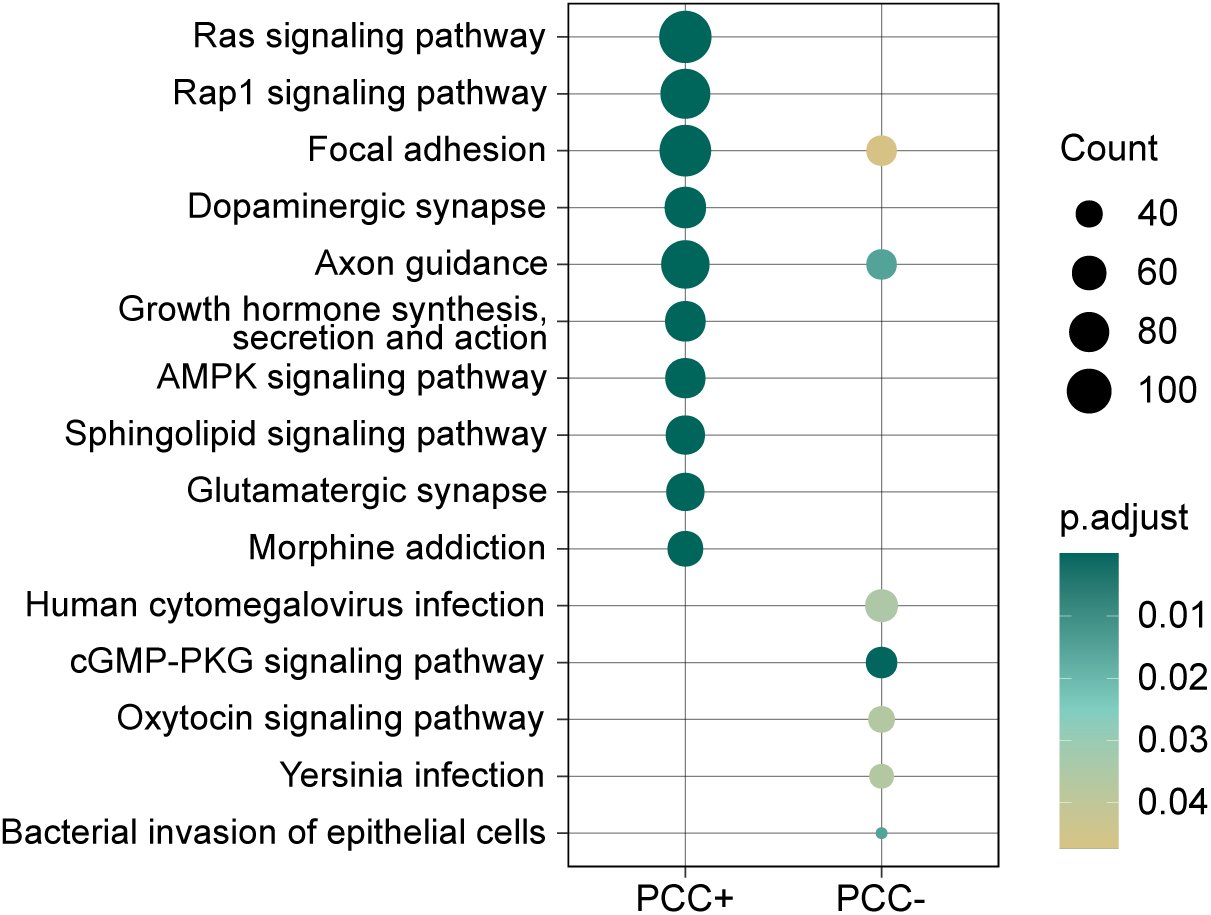
KEGG pathway enrichment analysis of differentially methylated genes (DMGs) between 3 and 12 months in individuals with PCC+ and PCC-. The horizontal axis represents different groups, while the vertical axis lists the enriched KEGG pathways. Node size indicates the number of DMGs associated with each pathway, and node color intensity corresponds to the adjusted p-value, with dark green colors reflecting more significant enrichment.

We complemented differential methylation analysis by identifying differential methylation regions (DMRs). Figure 2b) shows that the number of unique genes covered by significant DMRs decreased over time. KEGG pathway analysis of genes in DMRs at 3 months identified only one pathway; the AMPK signaling pathway. No significant KEGG pathways were identified at later time points using DMRs. Both in the analysis of DMCs and DMRs, we identified CPLX1, EID3, and TXNRD1 as enriched across all time points.

During the first year after infection, regression analysis identified CpGs with significantly associated with symptoms (Table 3). TXNRD1 was linked to neurological symptoms, specifically difficulties finding words, impaired memory function, and fatigue.

**Table 3:**
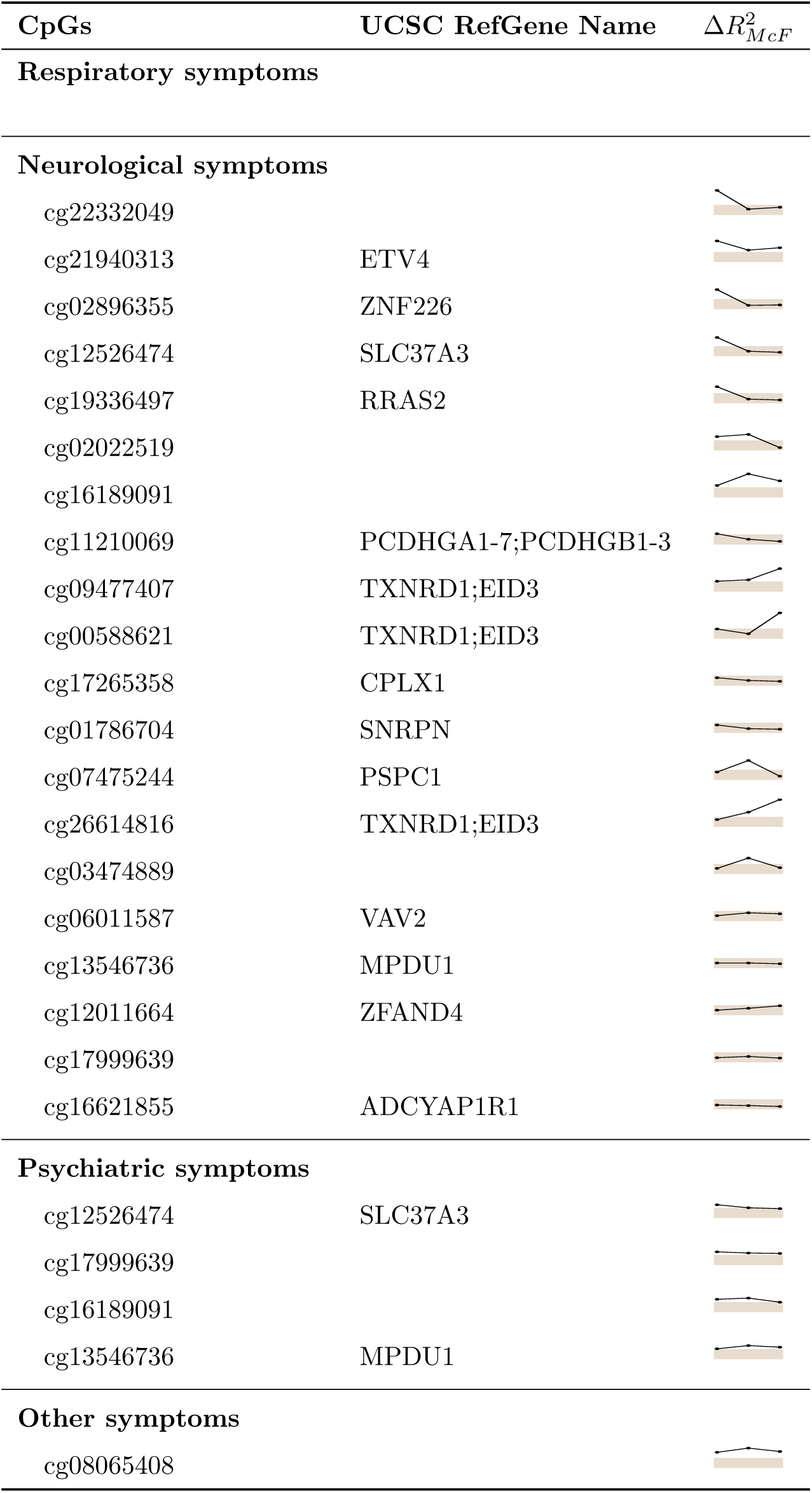
CpGs significantly associated with the symptoms at each time point (3, 6, and 12 months). The second column shows the names of genes associated with the CpGs. There are several CpGs that lack annotated genes. The third column shows sparklines representing the temporal trends of ΔMcFadden’s pseudo-R^2^ values 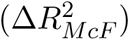 at 3, 6, and 12 months for each CpG. Each sparkline has three points corresponding to the 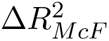 at these time points. The shaded area indicates 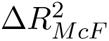 between 5% and 10%.

| CpGs | UCSC RefGene Name | $\Delta R^2_{McF}$ |
| --- | --- | --- |
| **Respiratory symptoms** |  |  |
| **Neurological symptoms** |  |  |
| cg22332049 |  |  |
| cg21940313 | ETV4 |  |
| cg02896355 | ZNF226 |  |
| cg12526474 | SLC37A3 |  |
| cg19336497 | RRAS2 |  |
| cg02022519 |  |  |
| cg16189091 |  |  |
| cg11210069 | PCDHGA1-7;PCDHGB1-3 |  |
| cg09477407 | TXNRD1;EID3 |  |
| cg00588621 | TXNRD1;EID3 |  |
| cg17265358 | CPLX1 |  |
| cg01786704 | SNRPN |  |
| cg07475244 | PSPC1 |  |
| cg26614816 | TXNRD1;EID3 |  |
| cg03474889 |  |  |
| cg06011587 | VAV2 |  |
| cg13546736 | MPDU1 |  |
| cg12011664 | ZFAND4 |  |
| cg17999639 |  |  |
| cg16621855 | ADCYAP1R1 |  |
| **Psychiatric symptoms** |  |  |
| cg12526474 | SLC37A3 |  |
| cg17999639 |  |  |
| cg16189091 |  |  |
| cg13546736 | MPDU1 |  |
| **Other symptoms** |  |  |
| cg08065408 |  |  |

Cell deconvolution applied to the DNAm data revealed no significant difference between PCC+ and PCC- (Supplementary Fig. A2). However, we observed a significant temporal decrease in the fraction of neutrophils over time (p-value *<* 0.05, ANOVA).

### 2.3 Differentially methylated regions undergo temporal changes in individuals with PCC+, but not the PCC- group

To further investigate the progression of methylation patterns in individuals with PCC+, we analyzed temporal changes over the first year post-infection. We identified 6697 DMGs that exhibited changes between 3 months and 12 months in individuals with PCC+, compared to only 2568 DMGs in the PCC- group. KEGG pathway analysis revealed several enriched pathways, with the Ras signaling pathway being the most significantly affected in individuals with PCC+, containing 111 DMGs.

Additional analysis of longitudinal changes in DMRs between 3 months and 12 months revealed 739 DMR-covered genes in the PCC+ group, while the PCC- group showed no significant DMRs. KEGG pathway enrichment analysis identified the PI3K-Akt signaling pathway and cytoskeleton regulation in muscle cells as the most enriched pathways (Supplementary Fig. A4).

### 2.4 No accelerated epigenetic aging in the PCC+ group relative to the PCC- group

Epigenetic aging refers to the accumulation of molecular changes in the DNA that reflect biological aging at the cellular level. Measuring the epigenetic age (EpiAge) acceleration, the deviation of epigenetic age from chronological age, has become a key indicator of aging-related conditions. Recent studies have reported accelerated EpiAge in individuals with COVID-19 and PCC compared to healthy controls [12, 13]. To investigate whether individuals with PCC+ in our cohort exhibit similar epigenetic aging patterns over time, we evaluated the differences between their EpiAge acceleration rates and those estimated for the PCC- group.

First, we measured the biological age of every sample at every measured time point (3, 6, and 12 months) using six blood-based DNAm age clocks (Horvath [14], Hannum [15], Horvath Skin and Blood [16], PhenoAge [17], Zhang [18], and NCAE-Age [19]), from which we calculated their EpiAge acceleration. We observed no statistically significant differences in EpiAge acceleration rates between the PCC+ and PCC- groups for any of the DNAm age clocks at any time point (Mann-Whitney U test, p-value *>* 0.05) (Fig. 4). Similarly, EpiAge acceleration rates obtained from the averaged EpiAge estimates between age clocks did not show significant differences between individuals with PCC+ and PCC-.

**Figure 4:**
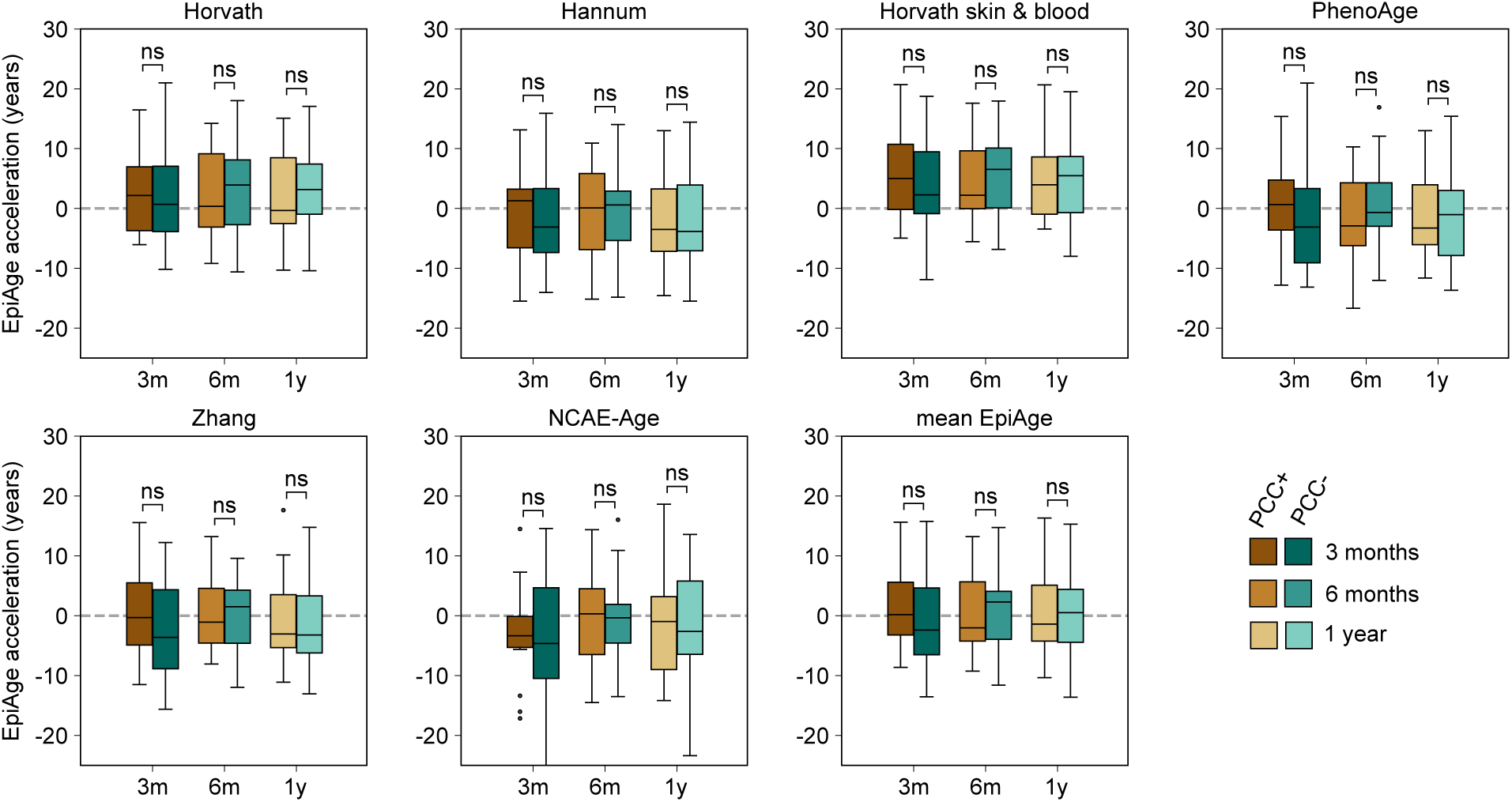
Epigenetic age acceleration of individuals with PCC+ and PCC- compared at 3, 6, and 12 months, from six DNA methylation age clocks plus their average estimation. Actual ages are adjusted by adding the corresponding number of years since baseline. The abbreviation “ns” denotes comparisons where no significant difference was observed (Mann-Whitney U test p-value *>* 0.05).

We expanded our analysis to investigate the correlation between the number of symptoms at each time point and EpiAge acceleration, aiming to understand the potential impact of symptom burden on biological aging. However, no significant correlation was found between symptom count and EpiAge acceleration, suggesting that symptom burden may not directly influence EpiAge acceleration in this context (Fig. A5).

## 3 Discussion

PCC manifests as an extensive range of symptoms, including problems with cognitive processing known as “brain-fog”, reduced lung function, autonomous nervous system impairment, chronic fatigue or anxiety, chronic cough, heart palpitations, and more [20, 4]. Most current studies have primarily studied hospitalized patients, and the duration of follow-up time has been relatively brief. Here, we have focused on long-term symptoms that lead to reduced health-related quality of life in a cohort of otherwise healthy adults monitored over one year after disease onset.

In this study, we identified distinct DNAm patterns that separate individuals with PCC+ and PCC-. The differences in DNAm profile between the PCC+ and PCC- groups diminish with time. We propose that this temporal change reflects the improvement many patients with PCC experience over time, as indicated by the decreasing number of reported symptoms observed at each follow-up in our study.

Common symptoms experienced by individuals with PCC include cognitive issues and fatigue. We observed that TXNRD1, which encodes Thioredoxin Reductase 1, was differentially methylated at each time point and showed an association with these symptoms. Thioredoxin Reductase 1 plays a critical role in the thioredoxin system, which is essential for reducing oxidative stress. This system is vital for maintaining cellular redox balance. Oxidative stress, along with mitochondrial dysfunction, has been implicated in PCC in numerous studies, suggesting a potential mechanism underlying these long-term symptoms [21, 22]. Al-Hakeim et al. (2023) reported that 32% of PCC patients in their cohort exhibited increased oxidative toxicity and reduced antioxidant defenses. Their findings suggest that neuropsychiatric symptoms following COVID-19 infection may have a neuro-oxidative origin [21]. We acknowledge that our results were obtained from PBMCs, while symptoms originate from other tissues. Given that COVID-19 is a systemic infection, we propose that oxidative stress occurs across multiple tissues post-infection, contributing to symptom development. Furthermore, using PBMC, we identify epigenetic modificaitons in pathways related to other organs (CNS, heart). As a matter of fact, it is well established that DNAm patterns found in blood cells can be reliable biomarkers for neurocognitive conditions [23]. The reason for these ubiquitous epigenetic changes remain enigmatic, but a study addressed DNAm alterations linked to autism spectrum disorder (ASD) and identified DNAm alterations that overlapped between blood and brain tissues of ASD patients [24].

Several mechanisms may contribute to a dysfunctional thioredoxin system and increased cellular oxidative stress. First, the main protease of SARS-CoV-2 has been shown to target TXNRD1 [25]. From a mechanistic perspective, this could align with a viral strategy to suppress DNA synthesis and conserve the ribonucleotide pool for enhanced virion production, which could lead to increased oxidative stress. Second, TXNRD1 is a selenoprotein, highly dependent on selenium [26]. Selenium deficiency has been linked to increased COVID-19 mortality and is proposed to elevate the risk of developing PCC [27].

KEGG pathway analysis of DMGs and DMRs at each time point revealed enrichment in several key signaling pathways. The most prominent pathways were AMPK (DMRs) and PI3K-Akt (DMGs). These findings align well with previous results from Bramante et al. (2023), who demonstrated that metformin reduces the incidence of PCC [28]. Metformin has previously been shown to reduce oxidative stress via the PI3K-Akt signaling pathway [29], as well as lowering intracellular reactive oxygen species levels and up-regulate thioredoxin expression through the AMPK signaling pathway [30]. Together, these insights provide a plausible mechanism for how metformin may reduce PCC symptom severity by modulating oxidative stress and key regulatory pathways involved in post-COVID recovery.

Recent research has explored additional roles of TXNRD1 besides its enzymatic role in cellular redox regulation. Hao et al. (2023) proposed TXNRD1 as a regulator of age-associated inflammation through its effects on the innate immune response [31]. Given the evidence of accelerated biological aging in COVID-19 patients and individuals with PCC, we aimed to investigate this further [12, 13]. Using a well-matched cohort of individuals with initial mild COVID-19 infection and low comorbidity burden, we applied various biological clocks based on DNAm data. However, we found no significant differences in epigenetic age acceleration between the PCC+ and PCC- groups at any time point.

Longitudinal analysis of DMGs over the first year after infection revealed significant enrichment of the RAS and RAP1 signaling pathways in KEGG pathway analysis, while DMR analysis highlighted the PI3K-AKT signaling pathway and cytoskeleton regulation in muscle cells. Our results suggest an early impact on these systems, as seen in the results from GO analysis at 3 months that reveal enrichment in small GTPase-mediated signal transduction as well as regulation of trans-synaptic signaling and modulation of chemical synaptic transmission. We also observed that genes associated with these pathways, such as VAV2 (Ras-related GTPase) and RRAS2 (a Ras-like low-molecular-weight GTPases), are associated at early time points with neurological symptoms but not as much in later time points. These early enrichments is in line with our previous report on COVID-19-induced DNAm changes mapping to the Ras-system [32] and suggest an early configuration of the RAS signaling pathway following infection.

RAS and RAP1 signaling pathways have previously been identified as enriched in the brains of deceased COVID-19 patients. Antunes et al. (2024) analyzed the brain proteome of fatal COVID-19 cases and compared it to differentially regulated proteins in post-mortem schizophrenia brains [33]. They reported that pathways specifically enriched after COVID-19 included Ras, Rap1, mitogen-activated protein kinases (MAPK), and immunological processes such as T-cell receptor and chemokine signaling pathways. Our findings further support the hypothesis that COVID-19 impacts these pathways during infection.

Small GTPases in RAS and RAP1 signaling pathways are central in many fundamental cell processes and connect multiple receptors and effectors inside cells throughout the body. RAS and RAP1 signaling pathways have previously been associated with multiple neurological disorders [34], and many commonly expressed symptoms in PCC might be explained by pathways connected to the RAS signaling pathway. For example, dysregulation of the RAP1-dependent, NO-mediated blood vessel relaxation could explain vascular symptoms such as chilblains and Raynaud’s phenomena [35]. Similarly, symptoms of sleep disturbance, nightmares, and cognitive symptoms in PCC may result from dysfunction of the NMDA receptor due to regulatory mechanisms of the RAS signaling pathway in synaptic plasticity at the NMDAR complex [36]. Longitudinal changes in DMGs in the dopaminergic and glutamatergic pathways support this hypothesis.

The precise connections and causal relationships between these pathways remain unclear based on our results alone, highlighting the need for further in vitro studies to confirm these findings. SARS-CoV-2 may disrupt redox homeostasis, potentially leading to dysfunction in small GTPase-regulated pathways, such as the RAS signaling system, given their sensitivity to oxidative stress. Alternatively, the virus might directly target these systems to facilitate cellular infection and replication, further exacerbating the observed dysregulation [37].

This study’s prospective, longitudinal design enabled the tracking of DNAm changes and symptom dynamics across multiple time points, offering insights into the mechanisms and progression of PCC+. By using matched groups with low levels of comorbidities and only mild initial COVID-19 infection, we reduced interfering factors in our analyses, enhancing our capacity to identify and analyze differentially methylated pathways in this cellular compartment. This approach allowed us to focus more directly on pathways potentially implicated in PCC without interference from unrelated comorbidities. The study would have benefited from larger cohort sizes, which would enhance the generalizability of our findings. Expanding the sample size could provide a more robust dataset, enabling stronger statistical power and potentially uncovering additional insights into methylation changes in PCC+. While the study suggests potential mechanisms behind PCC+, additional experimental or interventional studies are needed for validation.

## 4 Conclusion

This longitudinal study reveals dynamic DNAm patterns and potential mechanisms underlying long-term symptoms associated with PCC in PBMCs. By tracking methylation changes over one year in a well-matched cohort with mild initial COVID-19 infection, we identified specific methylation patterns that distinguished PCC+ individuals from COVID-19 convalescents. These differences in methylation profiles diminished over time, reflecting the gradual symptom improvement reported by many PCC+ patients.

Our analysis linked PCC symptoms – particularly cognitive impairment and fatigue – to methylation changes in TXNRD1, suggesting a connection to redox imbalance. Although we measured the methylome fallout in peripheral lymphocytes, these alterations likely reflect a broader systemic redox mismatch. Related pathways, such as PI3K-Akt and AMPK, were significantly enriched. These findings offer a potential explanation for the mechanism by which metformin reduces the incidence of PCC, shedding light on how it may influence key pathways involved in post-COVID symptoms.

While the study’s longitudinal design reduced the influence of confounding factors, larger cohort sizes would strengthen the generalizability of our findings. Future research should aim to confirm causality through experimental studies, potentially opening avenues for targeted therapeutic interventions.

## 5 Methods

### 5.1 Study design and study cohort

The study cohort consisted of patients from a prospective multicenter cohort study (CoVUm, clinical-trials.gov ID: NCT04368013), previously described in published papers [4]. PCR-confirmed, SARS-Cov-2 positive individuals were enrolled between April 2020 and June 2021. Both hospitalized (*≥* 18 years of age) and non-hospitalized (*≥* 15 years) patients were eligible for enrolment. Individuals who were unable to provide informed consent or unable to read or communicate in Swedish were excluded. Out of the 579 included individuals, we selected a non-hospitalized sub-cohort of 22 individuals with PCC+ and 22 COVID-19 convalescents (PCC-) from the same cohort, matched for sex, age, and BMI. Narrow exclusion criteria were used to reduce the impact on epigenetic expression from other variables. The individuals were selected without cardiovascular disease (not hypertonia), malignant disease, autoimmune diseases (not hypothyroidism), active smoking, COPD, liver disease, kidney diseases, or HIV. All included individuals had a mild initial infection according to the WHO clinical progression scale [38].

REDCap electronic data capture tools were used to enter participant data into electronic case report forms hosted by Umeå University [39]. Baseline characteristics, disease severity, level of care, and clinical parameters were collected at inclusion. The Charlson Comorbidities Index was used to quantify comorbidity-based disease burden and mortality risk [40]. Wilcoxon rank sum test was used to compare continuous variables between groups and Fisheŕs exact test for categorical variables.

### 5.2 Outcome measures

A custom questionnaire including 15 symptoms was used to assess persistent symptoms at every follow-up visit. The modified Medical Research Council scale was used to evaluate Self-experienced dyspnoea, and the cut-off for dyspnoea was set at score *≥* 1 [41].

Health-related quality of life after COVID-19 infection was assessed with the EuroQol 5-dimension 5-level questionnaire (EQ-5D-5L) and the EuroQol Visual Analogue Scale (EQ-VAS). The results were converted to an index value (EQ-5D Index) by using the United Kingdom as the reference population [42].

### 5.3 Post COVID-19 condition plus (PCC+) definition

PCC+ was, as in previous papers, defined as the prevalence of *≥* 1 symptom after 6-month follow-up, together with either moderate (score *≥* 3) difficulties in *≥* 2 dimensions of EQ-5D-5L and/or self-assessed overall health *≤* 60 in EQ-VAS. Individuals included met the definition at the 6- or 12-month follow-up or both [4].

### 5.4 Epigenome-wide DNAm analyses

PBMCs were isolated and DNA was prepared as described in [32]. Briefly, DNA was extracted from PBMCs using QIAamp DNA Mini Kit on the QIAcube Connect platform (Qiagen, 51304, 9002864, Germany). DNA quantity was measured with Qubit HS dsDNA (Q32851, invitrogen, USA) and the quality was evaluated with NanoDrop Spectrophotometer (ND-1000, Thermo Fisher Scientific, USA). The methylation array data was generated by Clinical Genomics Linköping, Linköping University, Sweden. For each sample, 250 ng of DNA extracted from PBMCs was subjected to bisulfite conversion using EZ DNA Methylation Kit (Zymo Research) and further processed according to the Illumina Infinium MethylationEPIC v1.0 (850K) protocol. The BeadChip arrays were scanned on the NextSeq 550 (Illumina) instrument.

### 5.5 Data pre-processing

Samples collected at 3 and 12 months were DNAm profiled together, while those collected at 6 months were profiled separately. To reduce technical variation without destroying the biological signal, we pre-processed the data independently for each profiling batch.

We read raw intensity (IDAT) files containing the methylated (Meth) and unmethylated (Unmeth) measurements using the R package minfi [43], and normalized the data using the R package ENmix [44]. For the best possible consistency, we used the recommended ENmix methods out-of-band (OOB) background estimation, REgression on Logarithm of Internal Control probes (RELIC) dye-bias correction, no quantile normalization, and the Regression on Correlated Probes (RCP) probe-type bias correction [45]. We represented the methylation level for each probe per sample by M-values, calculated as 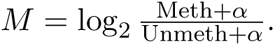

Next, we filtered out non-CpG probes, probes associated with SNPs, multi-hit probes, and probes located on the X or Y chromosomes using the R package ChAMP [46]. Additional batch corrections using the R function ComBat addressed slide and array effects [47]. Finally, we inferred cell type proportions in the samples using the R package EpiDISH [48].

### 5.6 Differential methylation analysis

We identified differentially methylated CpGs between PCC+ and PCC- groups using the R package limma [49], with age, sex, and cell type proportions included as covariates in the model. The longitudinal analysis focused on changes between 3 and 12 months, as samples collected at 6 months were DNAm profiled separately, making it challenging to distinguish technical noise from biological variation. To account for random effects in the longitudinal model, we included patient ID as a blocking factor when fitting the linear model to the data.

We classified CpGs as differentially methylated (DMCs) when the p-value was less than 0.01. After identifying DMCs, we mapped them to their corresponding differentially methylated genes (DMGs) using the function getFlatAnnotation from the R package missMethyl [50]. We considered a gene differentially methylated if it contained at least one DMC.

We identified differentially methylated regions (DMRs) using the R package DMRcate [51].

### 5.7 Disease pathways analysis

To analyze KEGG pathway enrichment, we mapped the identified DMGs to 334 KEGG pathways using the function enrichKEGG from the R package clusterProfiler [52], and visualized the top 10 enriched KEGG pathways with a Benjamini-Hochberg FDR-adjusted p-value less than 0.05.

### 5.8 Modelling symptom outcome on DNAm

To investigate the association between the methylation levels of specific CpGs and symptom clusters over the three time points (3, 6, and 12 months), we performed logistic regression using the R package glm. To make the analysis computationally feasible, we focused on 155 CpGs that demonstrated differential methylation consistently across the 3- and 6-month time points. This approach prioritized CpGs that showed early, since persistent changes are likely to be biologically relevant to symptom development. For each of these 155 CpGs, we constructed logistic models with symptom counts as the outcome and CpG beta values as predictors, fitting separate models for respiratory, neurological, psychiatric, and other symptom types.

Since symptoms can be influenced by the combined effects of multiple CpGs, we explored models that included subsets of CpGs. Given the limited sample size, we restricted these models to a maximum of two CpGs (referred to as saturated models) to minimize overfitting. All possible combinations of CpG pairs were evaluated, 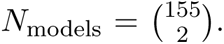 For each model, we estimated the variance in symptom occurrence explained by the included CpGs using McFadden’s pseudo-R^2^ 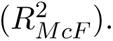. To identify the most explanatory CpGs, we used two criteria. First, we evaluated statistical evidence for each CpG by comparing the Akaike Information Criterion corrected (AICc) between models. Specifically, a focal CpG was considered supported if the AICc of the model with only that CpG was *≥* 10 units higher than the AICc of the saturated model [53]. Second, we approximated the relative importance of each CpG by calculating the difference in 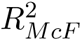 between the saturated model and the model, excluding the focal CpG. For each CpG, the median of these 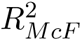 values was used to represent its relative importance. To ensure consistency across time, we filtered CpGs using a minimum explanatory threshold (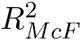 *>* 0.05) and included only those with significant associations at all three time points in the summary table.

### 5.9 Analysis of epigenetic age acceleration

Estimates of biological or epigenetic age (EpiAge) for individuals with PCC+ and PCC- were calculated using six blood-based DNAm age clocks, namely Horvath [14], Hannum [15], Horvath Skin and Blood [16], PhenoAge [17], Zhang [18], and NCAE-Age [19]. In addition, the mean predicted age across clocks was included to provide a robust and comprehensive estimate of biological age, minimizing clock-specific variability and sensitivity to outliers. DNAm age clocks were retrained, where applicable, for improved generalizability using 17,726 whole blood samples from healthy controls publicly available in the Gene Expression Omnibus (GEO) repository (accessed June 2023). EpiAge acceleration was measured as the deviation between the biological and chronological age of an individual. Actual ages were adjusted by adding the corresponding number of years since baseline. The statistical significance of the differences in EpiAge acceleration between PCC+ and PCC- individuals was assessed using the Mann-Whitney U test implemented in the SciPy library.

## 6 Ethical considerations

The study was approved by the Swedish Ethical Review Authority (approval number: 2020-01557) and was carried out according to the declaration of Helsinki.

## 7 Data Availability Statement

The datasets presented in this article are not readily available because ethical approval does not support publication of the entire dataset of the study cohort. Requests to access the datasets should be directed to.

## 8 Funding

The study was supported by grants from SciLifeLab, National COVID-19 Research Program (VC-2020-0015), financed by the Knut and Alice Wallenberg Foundation, Swedish Heart-Lung Foundation (20200325, 202100789 and 20220325).

## 9 Acknowledgements

We thank Sofie Degerman for her helpful suggestions on the data pre-processing strategies and Rubén Bernardo Madrid for his assistance with the methods for explaining the symptom associations.

## Appendix

**Figure A1:**
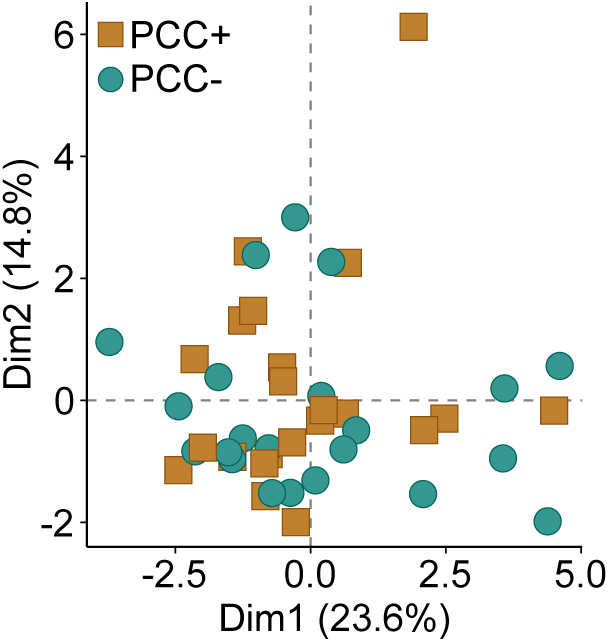
Principal component analysis of clinical chemistry data from the first 30 days after infection. Variables: Prothrombin time(INR), C-reactive protein, aspartate aminotransferase, alanine aminotransferase, creatinine, erythrocyte sedimentation rate, leukocytes, hemoglobin, thrombocytes, neutrophils, lymphocytes, monocytes, eosinophils, basophils, partial thromboplastin time test, D-Dimer. Abbreviations: PCC+, Post COVID-19 condition plus; PCC-, Matched COVID-19 convalescents.

**Figure A2:**
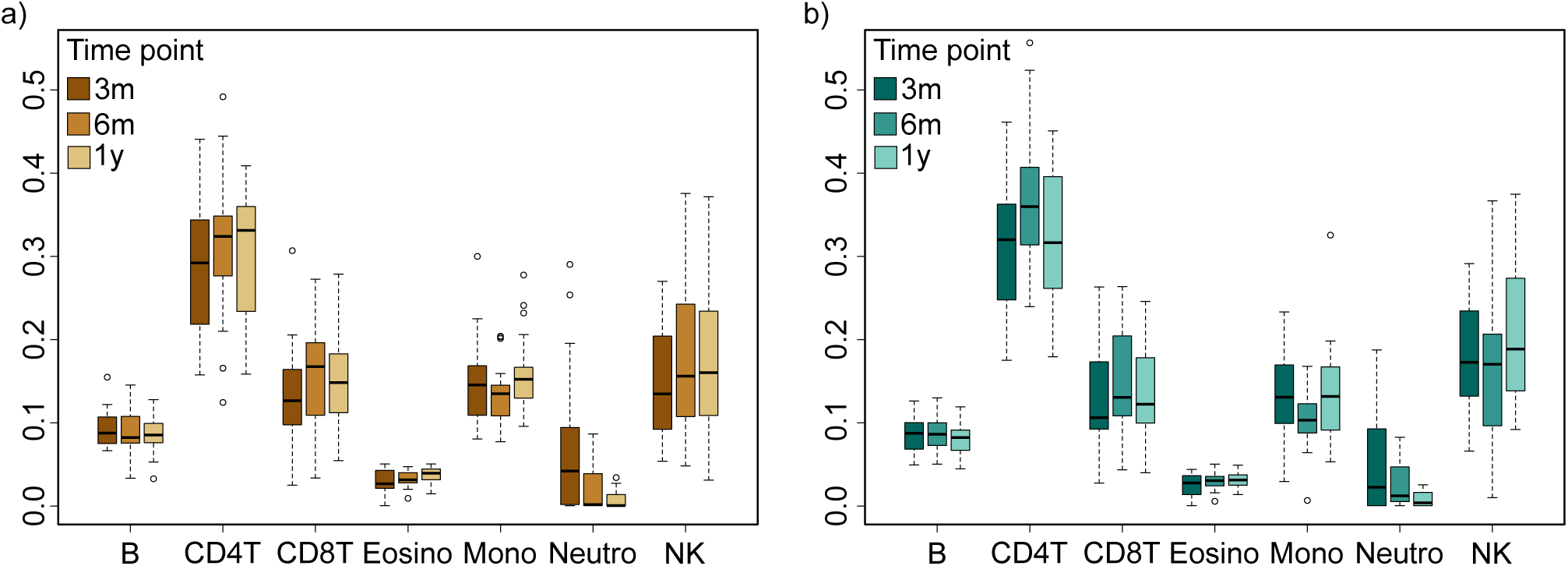
EPIDISH cell type analysis of a) PCC+ and b) PCC- groups. X-axis showing cell types including B-cells (B), CD4-positive T-cells (CD4T), CD8-positive T-cells (CD8T), eosinophils (Eosino), monocytes (Mono), neutrophils (Neutro) and natural killer T-cells (NK). Y-axis showing estimated cell-type fraction. PCC+ (Post COVID-19 condition plus), PCC- (Matched COVID-19 convalescents).

**Figure A3:**
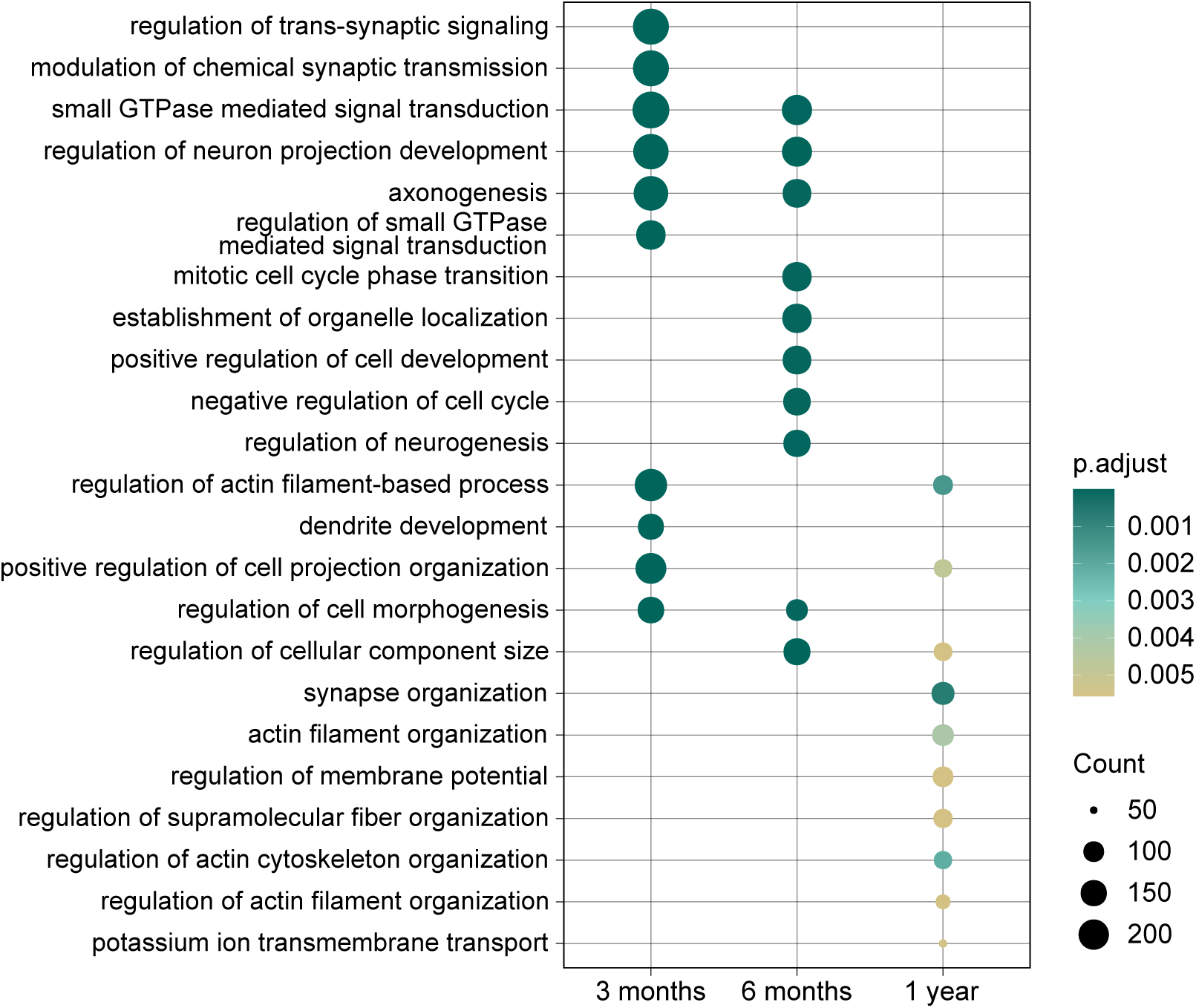
Gene Ontology (GO) enrichment analysis of differentially methylated genes (DMGs) between PCC+ (Post COVID-19 condition plus) and PCC- (matched COVID-19 convalescents) samples. The horizontal axis represents different time points, while the vertical axis lists the top 10 enriched GO pathways. Node size indicates the number of DMGs associated with each pathway, and node color intensity corresponds to the adjusted p-value, with red colors reflecting more significant enrichment.

**Figure A4:**
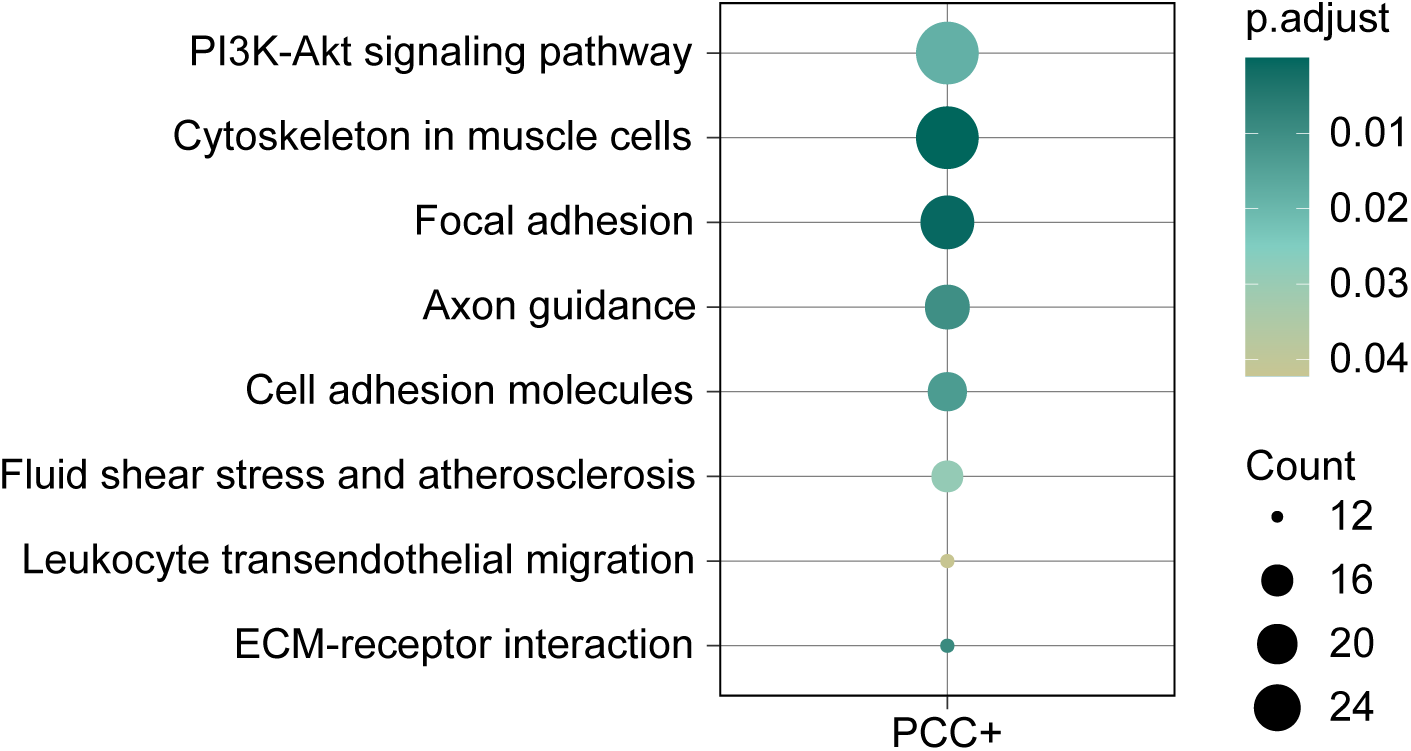
KEGG pathway enrichment analysis of differentially methylated regions (DMRs) between 3 and 12 months in individuals with PCC+ (Post COVID-19 condition plus). The vertical axis lists the enriched KEGG pathways. Node size indicates the number of genes in DMRs associated with each pathway, and node color intensity corresponds to the adjusted p-value, with red colors reflecting more significant enrichment.

**Figure A5:**
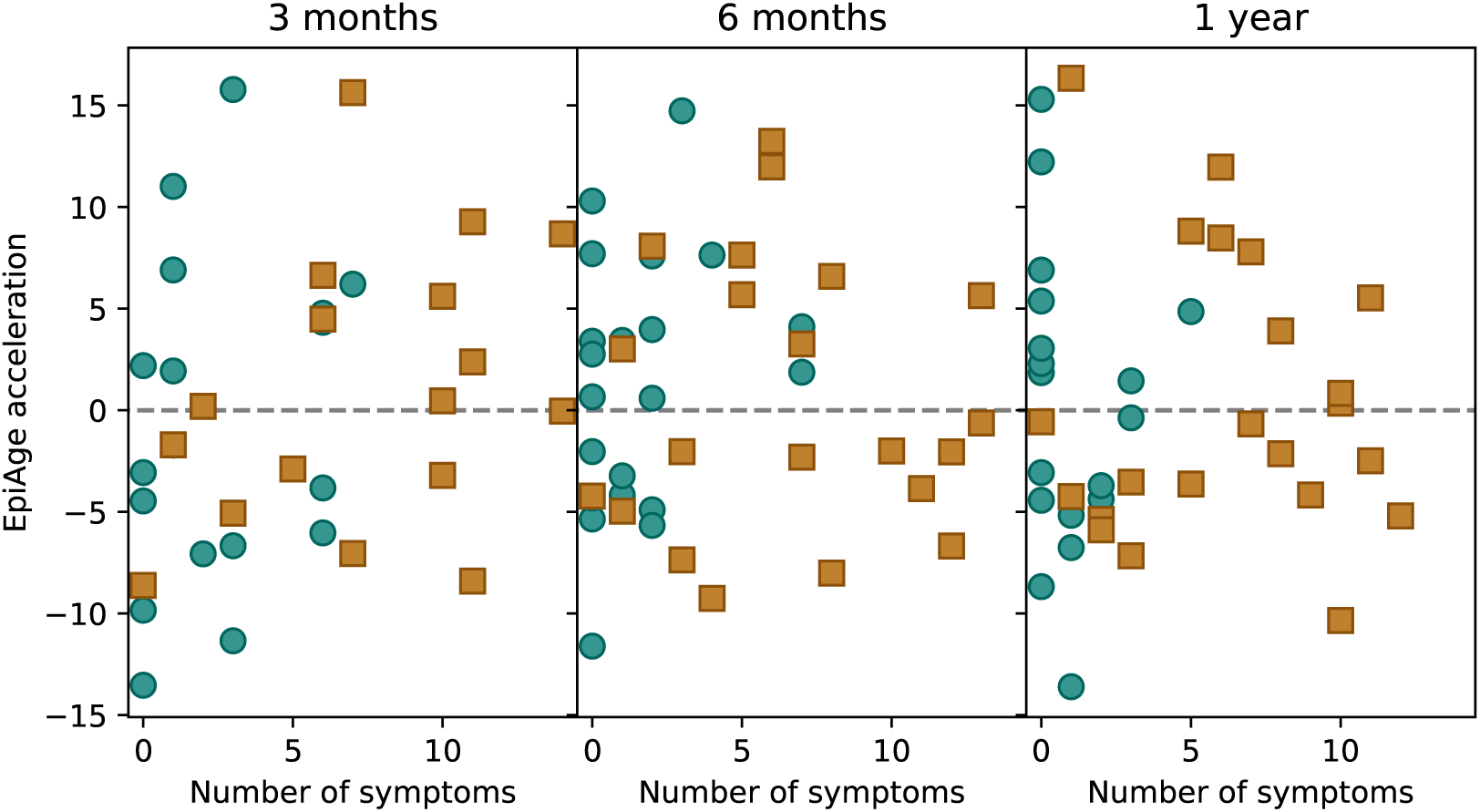
Correlation between number of symptoms and epigenetic age (EpiAge) acceleration at 3, 6 and 12 months after COVID-19 infection.

## Notes

### Competing Interest Statement

ML and MG are co-founders of PredictMe AB, a university spin-off company that develops DNA methylation data-based XAI disease risk classifiers.

### Author Declarations

The Swedish Ethical Review Authority gave ethical approval for this work (approval number: 2020-01557)

## References

[1] Hannah E Davis, Lisa McCorkell, Julia Moore Vogel, and Eric J Topol. Long covid: major findings, mechanisms and recommendations. Nature Reviews Microbiology, 2023.

[2] WHO. A clinical case definition of post covid-19 condition by a delphi consensus, 2021. https://www.who.int/publications/i/item/WHO-2019-nCoV-PostCOVID-19condition-Clinicalcasedefinition-2021.1.

[3] Lauren L. O’Mahoney, Ash Routen, Clare Gillies, Winifred Ekezie, Anneka Welford, Alexa Zhang, Urvi Karamchandani, Nikita Simms-Williams, Shabana Cassambai, Ashkon Ardavani, Thomas J. Wilkinson, Grace Hawthorne, Ffion Curtis, Andrew P. Kingsnorth, Abdullah Al-maqhawi, Thomas Ward, Daniel Ayoubkhani, Amitava Banerjee, Melanie Calvert, Roz Shafran, Terence Stephenson, Jonathan Sterne, Helen Ward, Rachael A. Evans, Francesco Zaccardi, Shaney Wright, and Kamlesh Khunti. The prevalence and long-term health effects of long covid among hospitalised and non-hospitalised populations: a systematic review and meta-analysis. eClinicalMedicine, Volume 55, 101762, 2022.

[4] Irma Ahmad, Alicia Edin, Christoffer Granvik, Lowa Kumm Persson, Staffan Tevell, Emeli Månsson, Anders Magnuson, Ingela Marklund, Ida-Lisa Persson, Anna Kauppi, Clas Ahlm, Mattias N. E. Forsell, Josefin Sundh, Anna Lange, Sara Cajander, and Johan Normark. High prevalence of persistent symptoms and reduced health-related quality of life 6 months after covid-19. Frontiers in Public Health, 11, 2023.

[5] Carlo Cervia-Hasler, Sarah C Brüningk, Tobias Hoch, Bowen Fan, Giulia Muzio, Ryan C Thompson, Laura Ceglarek, Roman Meledin, Patrick Westermann, Marc Emmenegger, et al. Persistent complement dysregulation with signs of thromboinflammation in active long covid. Science, 383(6680):eadg7942, 2024.

[6] Chansavath Phetsouphanh, David R Darley, Daniel B Wilson, Annett Howe, Cynthia Munier, Sheila K Patel, Jennifer A Juno, Louise M Burrell, Stephen J Kent, Gregory J Dore, et al. Immunological dysfunction persists for 8 months following initial mild-to-moderate sars-cov-2 infection. Nature immunology, 23(2):210–216, 2022.

[7] Shuk-Mei Ho, Abby Johnson, Pheruza Tarapore, Vinothini Janakiram, Xiang Zhang, and Yuet-Kin Leung. Environmental epigenetics and its implication on disease risk and health outcomes. ILAR journal, 53(3-4):289–305, 2012.

[8] Iain R Konigsberg, Bret Barnes, Monica Campbell, Elizabeth Davidson, Yingfei Zhen, Olivia Pallisard, Meher Preethi Boorgula, Corey Cox, Debmalya Nandy, Souvik Seal, et al. Host methylation predicts sars-cov-2 infection and clinical outcome. Communications medicine, 1(1):42, 2021.

[9] Frida Nikesjö, Shumaila Sayyab, Lovisa Karlsson, Eirini Apostolou, Anders Rosén, Kristofer Hedman, and Maria Lerm. Defining post-acute covid-19 syndrome (pacs) by an epigenetic biosignature in peripheral blood mononuclear cells. Clinical Epigenetics, 2022.

[10] Yunsung Lee, Espen Riskedal, Karl Trygve Kalleberg, Mette Istre, Andreas Lind, Fridtjof Lund-Johansen, Olaug Reiakvam, Arne V. L. Søraas, Jennifer R. Harris, John Arne Dahl, Cathrine L. Hadley, and Astanand Jugessur. Ewas of post-covid-19 patients shows methylation differences in the immune-response associated gene, ifi44l, three months after covid-19 infection. Scientific Reports, 2022.

[11] Joseph Balnis, Andy Madrid, Lisa A. Drake, Rachel Vancavage, Anupama Tiwari, Vraj J. Patel, Ramon Bossardi Ramos, John J. Schwarz, Recai Yucel, Harold A. Singer, Reid S. Alisch, and Ariel Jaitovich. Blood dna methylation in post-acute sequelae of covid-19 (pasc): a prospective cohort study. eBioMedicine, 2024.

[12] Luciano Calzari, Fernando Davide Dragani, Lucia Zanotti, Elvira Inglese, Romano Danesi, Rebecca Cavagnola, Alberto Brusati, Francesco Ranucci, Maria Anna Di Blasio, Luca Persani, Irene Campi, Sara De Martino, Antonella Farsetti, Veronica Barbi, Michela Zamperla Gottardi, Nicole Giulia Baldrighi, Carlo Gaetano, Gianfranco Parati, and Davide Gentilini. Epigenetic patterns, accelerated biological aging, and enhanced epigenetic drift detected 6 months following covid-19 infection: insights from a genome-wide dna methylation study. Clinical Epigenetics, 2024.

[13] Xue Cao, Wenjuan Li, Ting Wang, Dongzhi Ran, Veronica Davalos, Laura Planas-Serra, Aurora Pujol, Manel Esteller, Xiaolin Wang, and Huichuan Yu. Accelerated biological aging in covid-19 patients. Nature Communications, 2022.

[14] Steve Horvath. DNA methylation age of human tissues and cell types. Genome Biology, 14(10):R115, 2013.

[15] Gregory Hannum, Justin Guinney, Ling Zhao, Li Zhang, Guy Hughes, Srini Vas Sadda, Brandy Klotzle, Marina Bibikova, Jian Bing Fan, Yuan Gao, Rob Deconde, Menzies Chen, Indika Rajapakse, Stephen Friend, Trey Ideker, and Kang Zhang. Genome-wide Methylation Profiles Reveal Quantitative Views of Human Aging Rates. Molecular Cell, 49(2):359–367, 2013.

[16] Steve Horvath, Junko Oshima, George M. Martin, Ake T. Lu, Austin Quach, Howard Cohen, Sarah Felton, Mieko Matsuyama, Donna Lowe, Sylwia Kabacik, James G. Wilson, Alex P. Reiner, Anna Maierhofer, Julia Flunkert, Abraham Aviv, Lifang Hou, Andrea A. Baccarelli, Yun Li, James D. Stewart, Eric A. Whitsel, Luigi Ferrucci, Shigemi Matsuyama, and Kenneth Raj. Epigenetic clock for skin and blood cells applied to Hutchinson Gilford Progeria Syndrome and ex vivo studies. Aging, 10(7):1758–1775, jul 2018.

[17] Morgan E. Levine, Ake T. Lu, Austin Quach, Brian H. Chen, Themistocles L. Assimes, Stefania Bandinelli, Lifang Hou, Andrea A. Baccarelli, James D. Stewart, Yun Li, Eric A. Whitsel, James G. Wilson, Alex P. Reiner1, Abraham Aviv1, Kurt Lohman, Yongmei Liu, Luigi Ferrucci, and Steve Horvath. An epigenetic biomarker of aging for lifespan and healthspan. Aging, 10(4):573–591, 2018.

[18] Qian Zhang, Costanza L. Vallerga, Rosie M. Walker, Tian Lin, Anjali K. Henders, Grant W. Montgomery, Ji He, Dongsheng Fan, Javed Fowdar, Martin Kennedy, Toni Pitcher, John Pearson, Glenda Halliday, John B. Kwok, Ian Hickie, Simon Lewis, Tim Anderson, Peter A. Silburn, George D. Mellick, Sarah E. Harris, Paul Redmond, Alison D. Murray, David J. Porteous, Christopher S. Haley, Kathryn L. Evans, Andrew M. McIntosh, Jian Yang, Jacob Gratten, Riccardo E. Marioni, Naomi R. Wray, Ian J. Deary, Allan F. McRae, and Peter M. Visscher. Improved precision of epigenetic clock estimates across tissues and its implication for biological ageing. Genome Medicine, 11(1), 2019.

[19] David Martínez-Enguita, Sanjiv K. Dwivedi, Rebecka Jörnsten, and Mika Gustafsson. NCAE: data-driven representations using a deep network-coherent DNA methylation autoencoder identify robust disease and risk factor signatures. Briefings in Bioinformatics, 24(5), sep 2023.

[20] Thomas Bahmer, Christoph Borzikowsky, Wolfgang Lieb, Anna Horn, Lilian Krist, Julia Fricke, Carmen Scheibenbogen, Klaus F. Rabe, Walter Maetzler, Corina Maetzler, Martin Laudien, Derk Frank, Sabrina Ballhausen, Anne Hermes, Olga Miljukov, Karl Georg Haeusler, Nour Eddine El Mokhtari, Martin Witzenrath, Jörg Janne Vehreschild, Dagmar Krefting, Daniel Pape, Felipe A. Montellano, Mirjam Kohls, Caroline Morbach, Stefan Störk, Jens-Peter Reese, Thomas Keil, Peter Heuschmann, Michael Krawczak, and Stefan Schreiber. Severity, predictors and clinical correlates of post-covid syndrome (pcs) in germany: A prospective, multi-centre, population-based cohort study. eClinicalMedicine, 51:101549, 2022.

[21] Hussein Kadhem Al-Hakeim, Haneen Tahseen Al-Rubaye, Dhurgham Shihab Al-Hadrawi, Abbas F. Almulla, and Michael Maes. Long-covid post-viral chronic fatigue and affective symptoms are associated with oxidative damage, lowered antioxidant defenses and inflammation: a proof of concept and mechanism study. Molecular Psychiatry, 2023.

[22] Laetitia Gay, Valérie Desquiret-Dumas, Nicolas Nagot, Clara Rapenne, Philippe Van de Perre, Pascal Reynier, and Jean-Pierre Molès. Long-term persistence of mitochondrial dysfunctions after viral infections and antiviral therapies: A review of mechanisms involved. Journal of medical virology, 2024.

[23] Janine M LaSalle. Dna methylation biomarkers of intellectual/developmental disability across the lifespan. Journal of Neurodevelopmental Disorders, 17:10, 2025.

[24] Shan V Andrews, Brooke Sheppard, Gayle C Windham, Laura A Schieve, Diana E Schendel, Lisa A Croen, Pankaj Chopra, Reid S Alisch, Craig J Newschaffer, Stephen T Warren, et al. Case-control meta-analysis of blood dna methylation and autism spectrum disorder. Molecular autism, 9:1–11, 2018.

[25] Ignacio A. Gallardo, Daniel A. Todd, Stella T. Lima, Jonathan R. Chekan, Norman H. Chiu, and Ethan Will Taylor. Sars-cov-2 main protease targets host selenoproteins and glutathione biosynthesis for knockdown via proteolysis, potentially disrupting the thioredoxin and glutaredoxin redox cycles. Antioxidants, 12(3), 2023.

[26] Anieli Golin, Alexey A. Tinkov, Michael Aschner, Marcelo Farina, and João Batista Teixeira da Rocha. Relationship between selenium status, selenoproteins and covid-19 and other inflammatory diseases: A critical review. Journal of Trace Elements in Medicine and Biology, 75:127099, 2023.

[27] Lutz Schomburg. Selenium deficiency in covid-19—a possible long-lasting toxic relationship. Nutrients, 14(2), 2022.

[28] Carolyn T Bramante, John B Buse, David M Liebovitz, Jacinda M Nicklas, Michael A Puskarich, Ken Cohen, Hrishikesh K Belani, Blake J Anderson, Jared D Huling, Christopher J Tignanelli, et al. Outpatient treatment of covid-19 and incidence of post-covid-19 condition over 10 months (covid-out): a multicentre, randomised, quadruple-blind, parallel-group, phase 3 trial. The Lancet Infectious Diseases, 23(10):1119–1129, 2023.

[29] Behzad Khallaghi, Fatemeh Safarian, Sanaz Nasoohi, Abolhassan Ahmadiani, and Leila Dargahi. Metformin-induced protection against oxidative stress is associated with akt/mtor restoration in pc12 cells. Life Sciences, 148:286–292, 2016.

[30] Xinguo Hou, Jun Song, Xiao-Nan Li, Lin Zhang, XingLi Wang, Li Chen, and Ying H. Shen. Metformin reduces intracellular reactive oxygen species levels by upregulating expression of the antioxidant thioredoxin via the ampk-foxo3 pathway. Biochemical and Biophysical Research Communications, 396(2):199–205, 2010.

[31] Xue Hao, Bo Zhao, Martina Towers, Liping Liao, Edgar Luzete Monteiro, Xin Xu, Christina Freeman, Hongzhuang Peng, Hsin-Yao Tang, Aaron Havas, Andrew V. Kossenkov, Shelley L. Berger, Peter D. Adams, David W. Speicher, David Schultz, Ronen Marmorstein, Kenneth S. Zaret, and Rugang Zhang. Txnrd1 drives the innate immune response in senescent cells with implications for age-associated inflammation. Nature Aging, 2024.

[32] Johanna Huoman, Shumaila Sayyab, Eirini Apostolou, Lovisa Karlsson, Lucas Porcile, Muhammad Rizwan, Sumit Sharma, Jyotirmoy Das, Anders Rosén, and Maria Lerm. Epigenetic rewiring of pathways related to odour perception in immune cells exposed to sars-cov-2 in vivo and in vitro. Epigenetics, 17(13):1875–1891, 2022. PMID: 35758003.

[33] André S. L. M. Antunes, Guilherme Reis-de Oliveira, and Daniel Martins-de Souza. Molecular overlaps of neurological manifestations of covid-19 and schizophrenia from a proteomic perspective. European Archives of Psychiatry and Clinical Neuroscience, 2024.

[34] Salvatore J 3rd Cherra and Reagan Lamb. Interactions between ras and rap signaling pathways during neurodevelopment in health and disease. Frontiers in molecular neuroscience, 2024.

[35] Ramoji Kosuru, Bandana Singh, Sribalaji Lakshmikanthan, Yoshinori Nishijima, Jeannette Vasquez-Vivar, David X Zhang, and Magdalena Chrzanowska. Distinct signaling functions of rap1 isoforms in no release from endothelium. Frontiers in cell and developmental biology, 2021.

[36] Xuelai Fan, Wu Yang Jin, and Yu Tian Wang. The nmda receptor complex: a multifunctional machine at the glutamatergic synapse. Frontiers in cellular neuroscience, 2014.

[37] Beibei Zhang, Shuli Li, Juntao Ding, Jingxia Guo, and Zhenghai Ma. Rap1b: A cytoskeletal regulator advantageous to viral infection. iScience, 27(10):111023, 2024.

[38] John C Marshall et al. A minimal common outcome measure set for covid-19 clinical research. The Lancet Infectious Diseases, 2020.

[39] Paul A. Harris, Robert Taylor, Robert Thielke, Jonathon Payne, Nathaniel Gonzalez, and Jose G. Conde. Research electronic data capture (redcap)—a metadata-driven methodology and workflow process for providing translational research informatics support. Journal of Biomedical Informatics, 42(2):377–381, 2009.

[40] Caroline Roffman, John Buchanan, and Garry Allison. Charlson comorbidities index. Journal of physiotherapy, 62(3), 2016.

[41] O Nishiyama, H Taniguchi, Y Kondoh, T Kimura, K Kato, K Kataoka, T Ogawa, F Watanabe, and S Arizono. A simple assessment of dyspnoea as a prognostic indicator in idiopathic pulmonary fibrosis. European Respiratory Journal, 36(5):1067–1072, 2010.

[42] EQ-5D-5L. Available online at: https://euroqol.org/information-and-support/euroqol-instruments/eq-5d-5l/.

[43] Martin J. Aryee, Andrew E. Jaffe, Hector Corrada-Bravo, Christine Ladd-Acosta, Andrew P. Feinberg, Kasper D. Hansen, and Rafael A. Irizarry. Minfi: a flexible and comprehensive bio-conductor package for the analysis of infinium dna methylation microarrays. Bioinformatics, 30(10):1363–1369, 01 2014.

[44] Zongli Xu, Liang Niu, Leping Li, and Jack A. Taylor. Enmix: a novel background correction method for illumina humanmethylation450 beadchip. Nucleic Acids Research, 44(3):e20–e20, 09 2015.

[45] Anil PS Ori, Ake T Lu, Steve Horvath, and Roel A Ophoff. Significant variation in the performance of dna methylation predictors across data preprocessing and normalization strategies. Genome Biology, 23(1):225, 2022.

[46] Yuan Tian, Tiffany J Morris, Amy P Webster, Zhen Yang, Stephan Beck, Andrew Feber, and Andrew E Teschendorff. Champ: updated methylation analysis pipeline for illumina beadchips. Bioinformatics, 33(24):3982–3984, 08 2017.

[47] W. Evan Johnson, Cheng Li, and Ariel Rabinovic. Adjusting batch effects in microarray expression data using empirical bayes methods. Biostatistics, 8(1):118–127, 04 2006.

[48] Shijie C Zheng, Charles E Breeze, Stephan Beck, and Andrew E Teschendorff. Identification of differentially methylated cell types in epigenome-wide association studies. Nature methods, 15(12):1059–1066, 2018.

[49] Matthew E. Ritchie, Belinda Phipson, Di Wu, Yifang Hu, Charity W. Law, Wei Shi, and Gordon K. Smyth. limma powers differential expression analyses for rna-sequencing and microarray studies. Nucleic Acids Research, 43(7):e47–e47, 01 2015.

[50] Belinda Phipson, Jovana Maksimovic, and Alicia Oshlack. missmethyl: an r package for analyzing data from illumina’s humanmethylation450 platform. Bioinformatics, 32(2):286–288, 09 2015.

[51] Timothy J Peters, Michael J Buckley, Aaron L Statham, Ruth Pidsley, Katherine Samaras, Reginald V Lord, Susan J Clark, and Peter L Molloy. De novo identification of differentially methylated regions in the human genome. Epigenetics & chromatin, 8:1–16, 2015.

[52] Tianzhi Wu, Erqiang Hu, Shuangbin Xu, Meijun Chen, Pingfan Guo, Zehan Dai, Tingze Feng, Lang Zhou, Wenli Tang, LI Zhan, et al. clusterprofiler 4.0: A universal enrichment tool for interpreting omics data. The innovation, 2(3), 2021.

[53] David R Anderson and Kenneth P Burnham. Avoiding pitfalls when using information-theoretic methods. The Journal of wildlife management, pages 912–918, 2002.

